# Quantifying Within-Household Tuberculosis Transmission: A Systematic Review and a Prospective Cohort Study

**DOI:** 10.1101/2025.03.14.25323897

**Authors:** Chuan-Chin Huang, Alicia E. Madden, Mercedes C. Becerra, Roger Calderon, Alexander L Chu, Carmen Contreras, Judith Jimenez, Leonid Lecca, Rosa Yataco, Qi Tan, Zibiao Zhang, Elena Jauregui, Megan B. Murray

## Abstract

Household-based studies are widely used to assess tuberculosis (TB) transmission and evaluate preventive strategies. These studies typically assume that household contacts (HHCs) who develop TB are infected by their index patient, but community-acquired infections may introduce misclassification, potentially biasing results. We aimed to quantify the extent of within-household TB transmission using genetic linkage data.

We first analyzed a prospective cohort study conducted in Lima, Peru, where we enrolled microbiologically confirmed TB index patients and their HHCs, following them for one year. We applied whole-genome sequencing (WGS) and 24-locus mycobacterial interspersed repetitive unit-variable number tandem repeat (MIRU-VNTR) genotyping to determine genetic relatedness between index-HHC pairs. We then conducted a systematic review of household TB transmission studies that applied genotyping methods to assess the proportion of genetically linked index-HHC pairs across diverse settings.

In Lima, we analyzed 175 index-HHC pairs with high-quality WGS data. We classified 62% as genetically linked, suggesting household transmission. Matching proportions were higher for secondary HHC cases (68%) than co-prevalent cases (52%). Our systematic review identified 13 studies across various epidemiological settings. Among statistically robust studies, household transmission predominated in moderate TB incidence settings (<250 cases per 100,000 person-years), with genetic linkage exceeding 68%. However, in high-burden settings, within-household transmission varied widely, likely due to community-acquired infections and methodological differences.

In summary, our findings suggest that in settings with ≤250 TB cases per 100,000 person-years, 20–35% of household TB cases may be misclassified due to community transmission, with lower misclassification among child and female contacts. The extent of this issue in high-burden settings remains unclear.

## Introduction

Household-based studies are often used to identify risk factors for tuberculosis (TB) transmission and evaluate the efficacy of TB preventive therapies (TPT). Typically, these studies define the earliest diagnosed TB patient within a household as the index patient and subsequently monitor their household contacts (HHCs) for incident TB disease. This design assumes that HHCs who develop TB disease are likely to have been infected by their respective index patients. However, this assumption may be challenged, as some HHCs likely acquire TB infections from community sources, introducing non-differential misclassification of TB outcomes (1–5).

In principle, such non-differential misclassification biases study results toward the null, underestimating the true magnitude of associations. Nevertheless, when unexpected or strong associations emerge, questions regarding the robustness and validity of the findings frequently arise. These concerns become particularly important in high TB-burden settings, where higher levels of community-based TB exposure may increase the potential for misclassification, thus weakening conclusions derived from household-based studies (6, 7).

To better understand the impact of community-acquired TB infections on household-based transmission studies, it is necessary to quantify the proportion of genetically linked index-HHC pairs, as these are likely the result of within-household transmission. In this study, we address this question in two ways. First, we assess the proportion of index-HHC patient pairs that are genetically linked using data from a previously conducted longitudinal household study we conducted in Lima, Peru. We then incorporate the findings from this study into a systematic review that includes studies from a variety of different settings that have applied molecular genotyping methods to index-HHC patient pairs. By synthesizing evidence across diverse epidemiological contexts and combining it with our primary data, we aim to rigorously evaluate the robustness and validity of household-based study designs across settings with different TB burdens.

## Method-Lima Household-Based Cohort Study

### Study Setting and Design

We conducted a prospective cohort study in a catchment area of Lima, Peru, covering 20 districts with approximately 3.3 million residents. The study design and methodology have been described previously (5). In brief, between September 2009 and August 2012, we enrolled incident index pulmonary tuberculosis (TB) patients aged 16 years or older from 106 participating health centres. We confirmed TB diagnosis through microbiological methods, including sputum smear and bacterial culture. Upon enrolment, we used structured questionnaires to collect sociodemographic and clinical data from index patients, including symptom duration. Within two weeks of an index patient’s TB diagnosis, we invited their household contacts (HHCs) to participate in the cohort study. Consenting HHCs underwent TB symptom screening and those with positive results were referred for further evaluation. We followed HHCs not diagnosed with TB at enrolment for one year, with scheduled visits at 2, 6, and 12 months. We classified HHCs diagnosed with TB within 14 days of enrolment as co-prevalent TB patients and those diagnosed after 14 days as secondary TB patients. Whenever possible, we collected sputum samples from HHC TB patients at enrolment or during follow-up.

### Genotyping, Whole-Genome Sequencing and Genetic Distance Analysis

*Mycobacterium tuberculosis* (Mtb) isolates from culture-positive index and HHC patients were genotyped using 24-locus mycobacterial interspersed repetitive unit–variable number tandem repeat (MIRU-VNTR) typing and sequenced using whole-genome sequencing (WGS) on the Illumina HiSeq platform. Read lengths ranged from 100 to 150 base pairs, with a minimum coverage of 20-fold. Raw WGS reads were mapped to the Mtb H37Rv reference genome using the BWA-MEM algorithm (8).We identified variants, including single nucleotide polymorphisms (SNPs) using an alignment-based approach (9, 10). Isolates with <95% genome mapping coverage were excluded. We considered nucleotide calls missing if the valid depth of coverage was <10% of the isolate’s mean coverage, the mean read mapping quality was <10, or the alternative allele frequency was <85%. Variants within the PE/PPE gene family and previously identified high-error regions were excluded (11, 12). Index-HHC pairs where at least one isolate showed evidence of mixed infection were excluded from the analysis (13).

### Assessment of Within-household Transmission for Index-HHC patient pairs

As a proxy for within-household transmission, we estimated the proportion of index-HHC patient pairs that were genetically linked by MIRU-VNTR genotyping and WGS genetic distance analysis. In the MIRU method, a genetically-linked transmission event was defined as an index-HHC pair with 23 or more matched loci out of 24. In the WGS method, a genetically-linked transmission event was defined as a genetic distance of 12 or fewer SNPs between Mtb isolates from the index-HHC pair (14). We first reported the proportion of all index-HHC patient pairs that were genetically linked. We then presented the matching proportions separately for index-co-prevalent and index-secondary HHC pairs. Given that many household-based studies restrict analyses to child contacts—who are more likely to be infected by their index patient than adult contacts—we also reported the matching proportions of pairs with a child contact separately (15–17). Finally, we evaluated the associations between index and HHC characteristics and within-household transmission among index-secondary HHC patient pairs using Fisher’s exact test based on WGS data.

## Results-Lima Household-Based Cohort Study

Among 12,676 household contacts (HHCs) exposed to a microbiologically confirmed TB index patient, 692 (5%) were diagnosed with TB at enrolment or during follow-up. Of these, 246 (36%) had culture-confirmed TB.

### MIRU genotyping matching

Among the culture-confirmed patients, 192 (78%) were successfully genotyped using MIRU-VNTR. Genotyping data for both the index and HHC patients were available for 168 pairs. Of these, 23 pairs (14%) had evidence of mixed infection and were excluded, leaving 145 pairs (86%) with high-quality MIRU data. Within this subset, 81 (56%) were genetically linked and classified as resulting from within-household transmission. Among the 51 index-co-prevalent HHC pairs, 26 (51%) were genetically linked, while among the 94 index-secondary HHC pairs, 54 (57%) were genetically linked. The HHC patients of 28 pairs were children. Of these, 5 were index-co-prevalent, of which 4 (80%) matched; 23 were index-secondary, of which 15 (65%) matched.

### WGS matching

In 190 index-HHC patient pairs with WGS data available for both the index and HHC patients, 15 pairs (5%) had evidence of mixed infection and were excluded, leaving 175 pairs (95%) with high-quality WGS data. Of these, 109 (62%) were genetically linked and classified as due to within-household transmission. Among the 175 pairs, we classified 66 (38%) as index-co-prevalent pairs and 109 (62%) as index-secondary pairs. We noted within-household transmission in 34 of 66 (52%) index-co-prevalent pairs and 74 of 109 (68%) index-secondary pairs. When restricting to the 33 pairs with a child HHC, we noted 5/8 (63%) index-co-prevalent pairs and 19/25 (76%) index-secondary pairs (Appendix Table A1). We also found a significantly higher matching proportion among pairs with female contacts (79%) compared to pairs with male contacts (54%; p=0.008) (Appendix Table A1).

## Method-Systematic Review

### Search Strategy and Study Selection

We searched PubMed, Web of Science, and Embase for papers that reported Mtb genotyping results for index and contact TB patients within the same household. We used search terms related to the household study design to find papers published between January 1^st^, 2006, and January 21^st^, 2025. These dates reflect the time period following the widespread adoption of standardized 12-locus MIRU-VNTR (18). Appendix Table A2 describes the search strategy for each database.

### Screening

We performed an initial title and abstract review, excluding papers that did not describe a study of the household contacts of index TB patients. Among those included, we searched full-text files for the following keywords related to the molecular typing of TB isolates: “WGS”, “whole genome sequencing”, “spoligotyping”, “spoligotype”, “spacer oligonucleotide typing”, “PCR”, “RFLP”, “restriction fragment length polymorphism”, “sequenced”, “typed”, “molecular typing”, “MIRU”, “VNTR”.

Publications containing one or more keywords underwent full-text evaluation by two independent reviewers; disagreements were resolved by discussion with a third reviewer. We excluded publications if they (a) did not involve original research on TB patients, (b) did not include information on household membership, (c) did not apply a qualifying genotyping method (spoligotyping, RFLP, MIRU-VNTR, whole-genome sequencing) to Mtb samples, (d) did not report the proportion of household index-contact pairs with matching Mtb genotyping, (e) enrolled fewer than five households, or (f) obtained genotyping results for fewer than five index-contact pairs. We examined the reference lists of relevant systematic reviews and meta-analyses for additional qualifying studies. A validation sample of 227 records was independently reviewed by a fourth reviewer, confirming the accuracy of the data extraction process.

### Data Extraction

For qualifying studies, two reviewers extracted the following data: location and dates of patient enrolment, study design, eligibility criteria for index patients and contacts, procedure for identifying HHC patients, definitions of household contacts, genotyping method(s), and the genotyping matching proportions for index and HHC patients. We initially intended to extract individual patient data to assess risk factors for genotype matching, but this information was not available for most studies. Discrepancies were resolved by a third reviewer. We extracted genotyping results based on a study’s most stringent definition of genotype matching; if genotyping results for individual patients were available, we calculated the proportion of patient pairs with identical genotypes. We reported the combined matching proportion for all co-prevalent and secondary HHC patients, then, when possible, reported matching among secondary HHC patients alone. We also calculated Clopper-Pearson exact 95% confidence intervals for the matching proportions. If study site-specific TB incidence was not reported, we used the WHO-reported national TB incidence estimate from the first year of patient enrolment or the year 2000, whichever was later (19). We also extracted individual-level metadata when available to evaluate the associations between patient characteristics and the likelihood of within-household transmission.

### Quality Assessment

We evaluated study quality based on study design rigor and statistical robustness. Study design rigor was assessed using an eight-question scoring framework (score range: 0–8), modified from a previous study, which focused on index patient enrolment, household definitions, HHC screening and follow-up, and genotyping completeness (Appendix Table A3). We evaluated statistical robustness using the width of the Clopper-Pearson exact 95% CIs for the matching proportion, considering intervals smaller than 40% to be robust.

## Result—Systematic Review

The study selection process is shown in Figure 1. Out of 2,532 unique citations identified from the database search, we did a full text review of 84 publications. In addition, colleagues alerted us to one publication meeting the study definition. Two reviewed studies from Brazil were follow-up publications from an earlier and larger genotyping study, so we reported data from the larger study (20). We identified one additional study from the citation list of a reviewed meta-analysis. Ultimately, thirteen articles met all inclusion and exclusion criteria (20–32).

**Figure 1.**
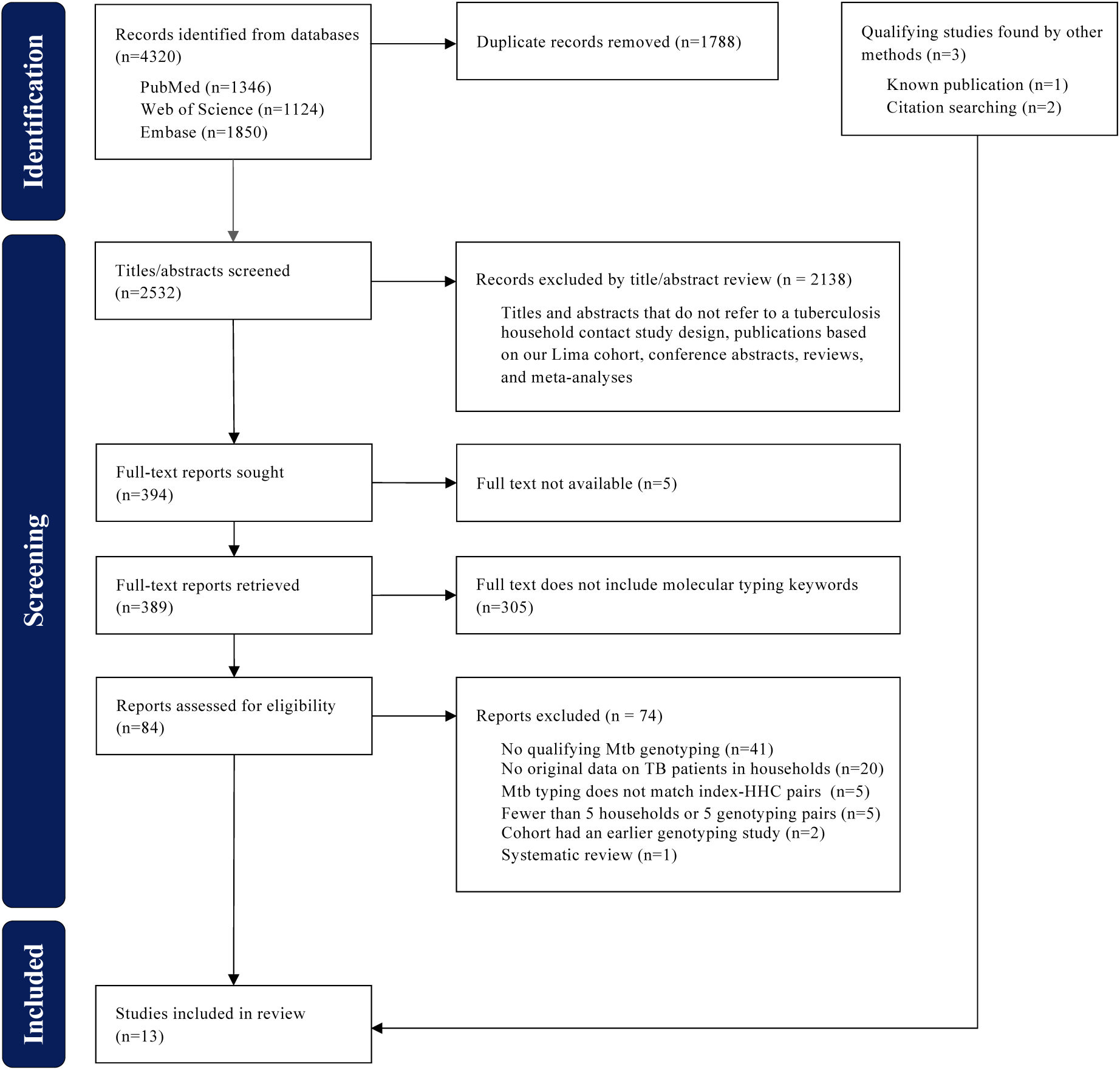
PRISMA Diagram of Study Selection Process.

The thirteen included studies, all published in English, evaluated within-household transmission between 731 household contacts and their index patients between 1995 and 2012 (Table 1). These studies were conducted in sub-Saharan Africa (South Africa, Uganda, Malawi), South America (Brazil, Argentina, Peru), central and Western Europe (Poland, Spain, England), Southeast Asia (Vietnam, Philippines), and Hong Kong. The studies encompassed a wide range of TB burden settings, with community incidence rates ranging from 14 to 742 TB patients per 100,000 person-years. The number of index-HHC pairs with genotyping or WGS data ranged from 6 to 344, with a median of 37 pairs. One study (21) included co-prevalent household patients only, five studies (20, 23, 24, 28, 31) included secondary household patients only, and seven studies (22, 25–27, 29, 30, 32) included both. Two studies restricted enrolment of index patients to MDR-TB patients (26, 27). Among the thirteen studies, age and gender information for index-HHC patient pairs were partially available for three studies and completely available for two studies (Appendix Tables A4 and A5).

**Table 1.**
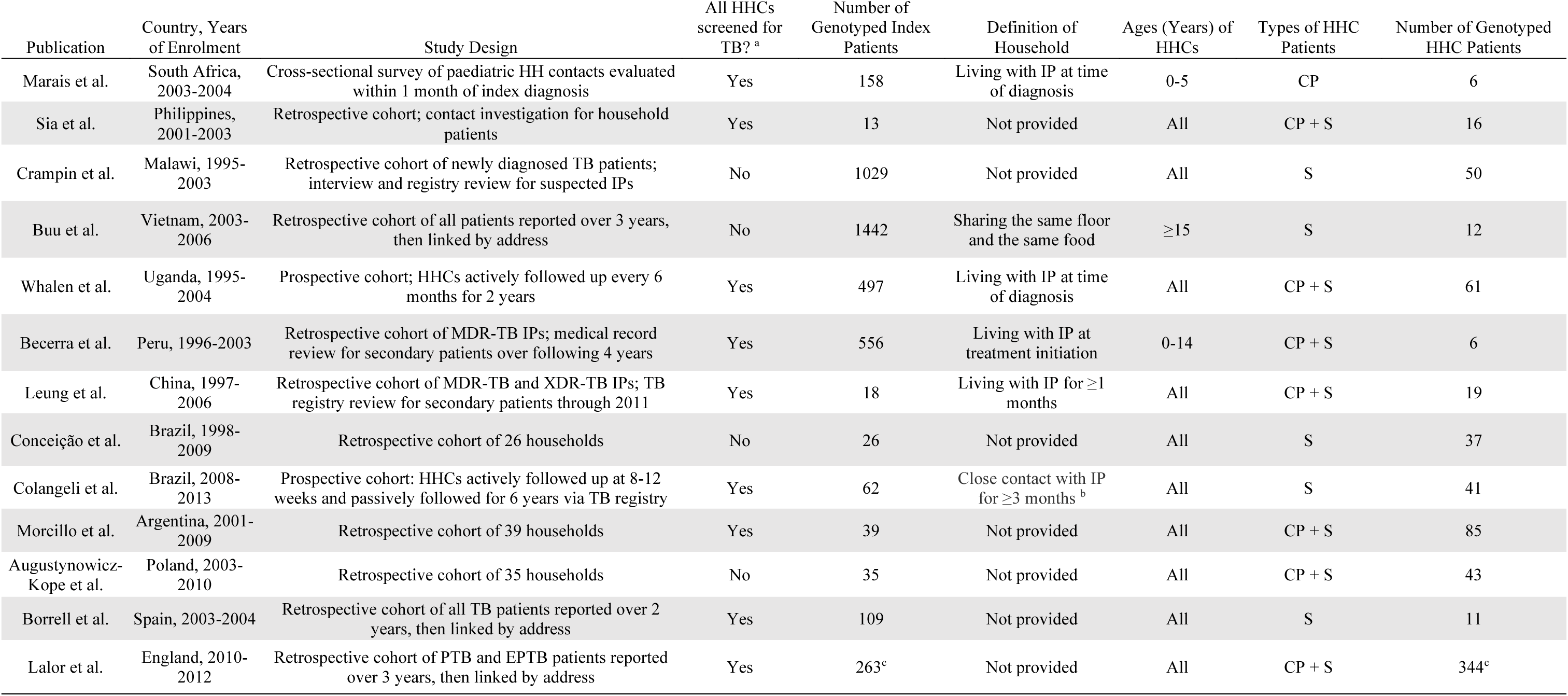
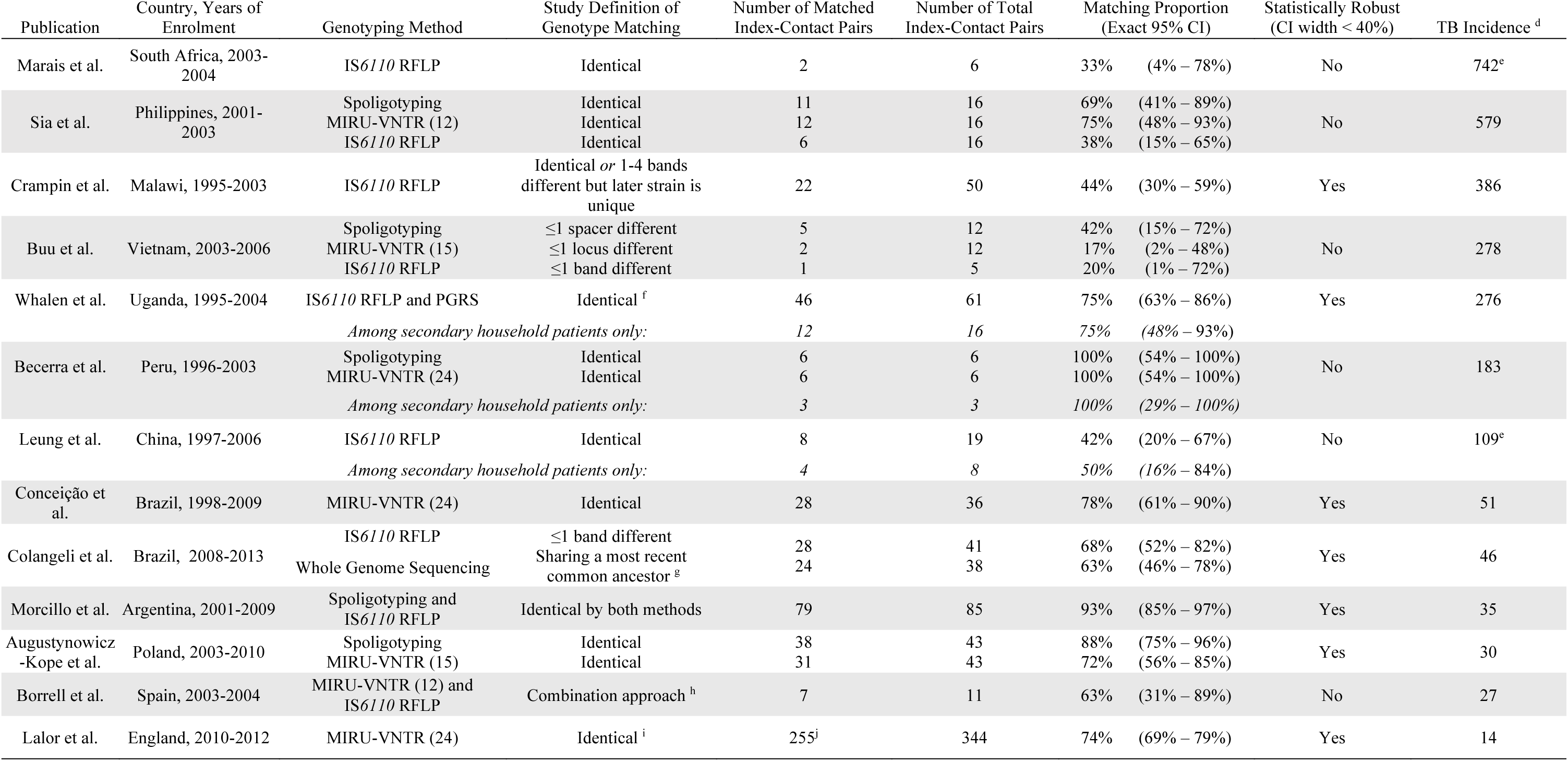
Characteristics of Qualifying Studies. Abbreviations: HHC, household contact; IP, index patient: CP, co-prevalent: S, secondary; (E)PTB, (extra-)pulmonary tuberculosis Abbreviations: MIRU-VNTR, mycobacterial interspersed repetitive-unit-variable-number tandem-repeats; RFLP, restriction fragment length polymorphism; PGRS, polymorphic GC-rich repetitive sequence ^a^ An initial screening of all HHCs was reported to have been performed by a research team or by the TB program. ^b^ Close contact defined as sleeping under the same roof ≥5 days/week, or sharing meals ≥5 days/week; or watching TV nights on weekends, or other significant contact (85% visited the household ≥18 days/month) ^c^ Estimated from 607 total genotyped TB patients based on study average of 2.3 patients per household ^d^ WHO estimated national incidence per 100,000 person-years, unless otherwise specified ^e^ Estimate of local TB incidence provided by the publication ^f^ More than five copies of IS6110 with identical fragments, or fewer than six copies of IS6110 with identical fragments and identical PGRS patterns ^g^ All matched pairs had fewer than 13 single nucleotide polymorphisms ^h^ RFLP patterns containing >6 IS6110 bands at the same positions, ≤6 IS6110 bands at the same positions but identical MIRU types, or a unique band difference in IS6110 patterns and identical MIRU12 types ^i^ Identical 24-locus MIRU-VNTR, or indistinguishable MIRU-VNTR where one or both patients were not typed to 23-loci ^j^ Estimated from 452 total confirmed and probable household transmission patients based on study average of 2.3 patients per household

### Quality Assessment

#### Study Design Rigor

Among the thirteen studies from the systematic review, all but three studies (28–30) had a score of five or higher. The most common gaps identified in the study design were the failure to report the proportion of eligible index patients who agreed to participate (Table A2, question 2), the lack of active follow-up to detect incident TB among HHCs (Table A2, question 7), and the prolonged interval allowed between the index patient’s diagnosis and the subsequent diagnosis of TB in HHC patients (Table A3, question 8).

#### Statistical Robustness

Among the thirteen studies, six (43%) reported matching proportions with a Clopper-Pearson exact 95% confidence interval width exceeding 40% (21, 22, 24, 26, 27, 31). Therefore, only seven reviewed studies were considered statistically robust (Figure 2).

**Figure 2.**
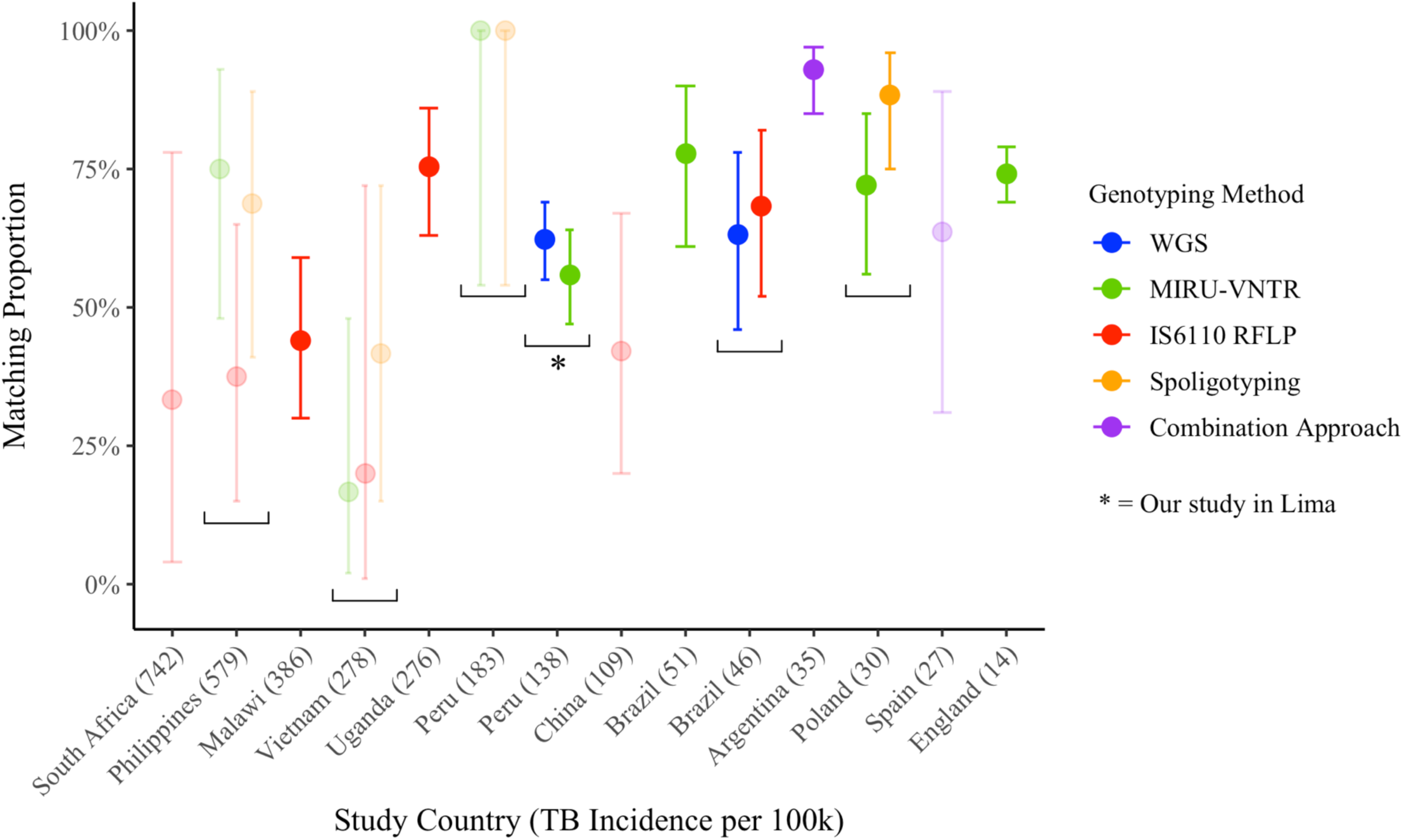
Genotyping Matching Proportions for All Studies and Genotyping Methods. Studies with confidence intervals smaller than 40% are represented with full opacity, while studies with confidence intervals greater than 40% are shown with transparency.

### Host Factors and Within-Household Transmission

Among the seven reviewed studies that met our criteria for statistical robustness, three included metadata on genetic linkage in both child and adult contacts (Table A3). However, two of these studies (28, 30) genotyped three or fewer total child contacts. In the third study (25), the matching proportion was 90% (26/29) in pairs with child contacts and 63% (20/32) in pairs with adult contacts. Two statistically robust studies reported gender information for comparison (Table A4). Both studies (28, 30) found a slightly higher matching proportion in pairs with female household contacts compared to those with male contacts (73% vs. 70% and 88% vs. 81%).

## Results—Proportion of Within-Household Transmission Among All Fourteen Studies

### All pairs (including index-co-prevalent and index-secondary patient pairs)

Across all fourteen studies (thirteen from the systematic review and one from our study in Lima, Peru), the proportion of index-HHC pairs that were genetically matched ranged from 17% to 100%. Among the six studies that employed multiple genotyping methods independently (i.e., the application of one method was not conditional on the results of another), five reported variability in the matching proportion across methods. While no genotyping method universally reported a higher or lower matching proportion compared to other methods, spoligotyping yielded the highest matching proportion in three out of four studies that used it. Only two studies implemented WGS as a tool for evaluating the matching proportion of index-HHC patient pairs (20). Eight studies used MIRU-VNTR (22, 24, 26, 28, 30–32), but the number of loci analysed varied (Table 1).

Among the eight studies with greater statistical robustness (20, 23, 25, 28–30, 32), seven reported a matching proportion ≥62%, with one study from Malawi reporting a proportion of 49%. All seven studies that reported a matching proportion ≥62% were conducted in settings with a TB incidence of ≤276 patients per 100,000 person-years (Figure 2), while the study from Malawi was conducted in an area with a TB burden of 386 patients per 100,000 person-years.

### Index-secondary patient pairs only

Ten studies, including our study in Lima, reported a separate matching proportion for index-secondary patient pairs (Figure 3). Of these, five studies had matching proportions with Clopper-Pearson 95% CIs ≤40% in width. Among these five studies, one conducted in Malawi (with a TB burden of 386 patients per 100,000 person-years) reported a matching proportion of 49%. The remaining four studies, all conducted in settings with a TB burden ≤138 patients per 100,000 person-years, reported matching proportions ≥68%.

**Figure 3.**
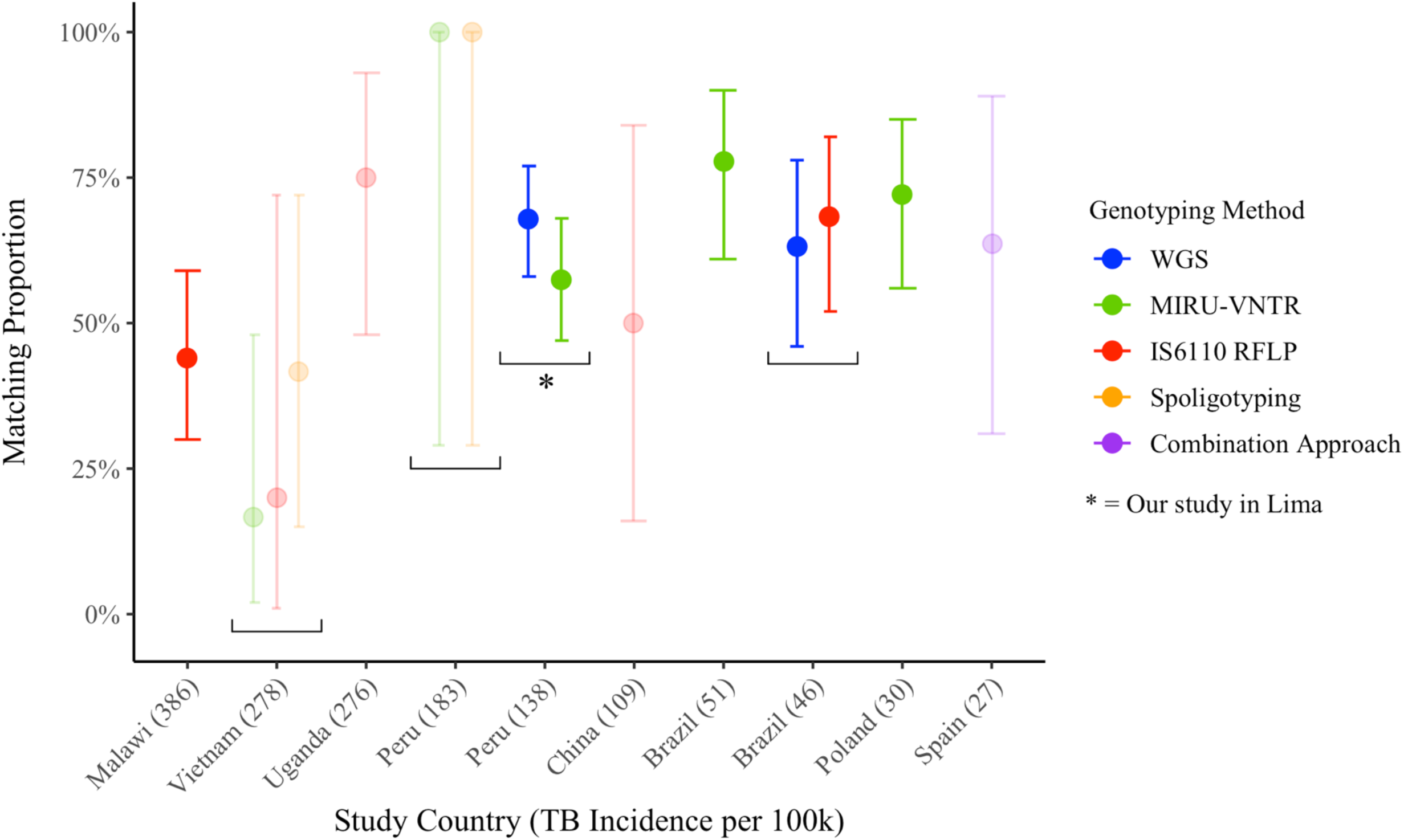
Genotyping Matching Proportions for Secondary Patient Pairs. Co-prevalent household contact patients are excluded. Studies with confidence intervals smaller than 40% are represented with full opacity, while studies with wide confidence intervals greater than 40% are show with transparency

## Discussion

Here, we found that in settings with TB incidence ≤250 cases per 100,000 person-years, the proportion of genetically matched index-secondary patient pairs consistently exceeds 68%, regardless of the genotyping method employed. This consistency indicates that among individuals exposed to TB at home, within-household transmission predominates in moderate-incidence settings. However, substantial variation was observed in settings with higher TB burdens (>250 cases per 100,000 person-years), variation that is likely due to limited sample sizes and genotyping data.

Genotyping tools differ in resolution, with WGS providing the highest resolution, RFLP and MIRU-VNTR offering moderate resolution, and spoligotyping the lowest. Higher-resolution methods generally yield lower genetic matching proportions (33). Within the same studies, matching proportions varied depending on the genotyping method used. While spoligotyping typically produced higher matching proportions, no single method among WGS, RFLP, or MIRU-VNTR consistently resulted in higher or lower proportions across studies. Several factors may explain these inconsistencies. WGS method, due to its high resolution, can detect mixed TB infections more frequently, leading to their exclusion from matching proportion calculations. Its higher sensitivity also makes it more susceptible to variations in sputum sample quality. Both factors can alter the numerators and denominators used to calculate matching proportions, contributing to discrepancies with RFLP and MIRU-VNTR.

Although all studies defined the earliest diagnosed TB patient in a household as the index case, only two actively and prospectively followed HHCs to track incident TB disease. Most studies relied on national TB registries to retrospectively identify secondary cases, often diagnosed years after the index patient. In the study from Malawi that reported a matching proportion of 44%, the time between index and contact patient diagnosis was allowed to be up to eight years. This delay increases the likelihood that household cases are community-acquired rather than originating from household transmission. If these retrospective studies had limited the diagnostic interval between index and secondary HHC patients to two years or less, reported matching proportions would likely have been higher. Thus, in settings with a TB incidence of ≤250 cases per 100,000 person-years, the actual matching proportion likely exceeds the reported 68%.

Household contacts’ age and gender may also influence within-household transmission dynamics. Many household-based studies, particularly clinical trials of TB preventive therapy, focus on child contacts to minimize misclassification, assuming that children have limited social interactions and are primarily infected by their index patient (5, 34, 35). Our findings support this approach, showing consistently higher matching proportions among child contacts than adults. Gender-based household roles may further shape transmission. Several systematic review studies indicate that women spend more time at home due to caregiving and domestic responsibilities (36–39), likely leading to increased exposure to infectious household members. This likely explains the higher matching proportions observed among female HHCs.

Our systematic review has limitations. Despite a rigorous search strategy, we may have inadvertently missed relevant studies due to search term constraints. Most included studies relied on data collected over a decade ago. Advances in diagnostics, community-based TB control, and expanded TPT coverage may have altered transmission dynamics, meaning older findings may not fully reflect current realities.

In summary, our findings suggest that in settings with a TB incidence of ≤250 cases per 100,000 person-years, 20–35% of incident TB cases among HHCs may be misclassified as household transmission. This misclassification is likely lower when analyses focus on child or female contacts. However, its impact in high-incidence settings remains unclear. TB preventive therapy (TPT) trials often span multiple countries with diverse TB burdens, requiring explicit consideration of misclassification differences across sites. Predictive modelling has been proposed to quantify and mitigate community-acquired infections in household studies (6, 7). Standardized methodologies and further validation of these models are needed to better characterize household transmission in high-burden settings.

## Data Availability

All data used in this study will be made available upon request.

## Supplementary Appendix

### Study Definition

We sought to identify all publications which met the following study description: a *Mycobacterium tuberculosis* (Mtb) genotyping study that reports genetic matching between index and household contact TB patients within the same household. Household contact patients could include both co-prevalent and secondary patients. Included studies analysed the Mtb strains from both index and household contact patients using a qualifying molecular typing technique and apply a reasonable definition of genetic matching. We included studies that also reported on non-household contacts if the study also reported a separate matching proportion for household contacts. We included studies that described the Mtb genotyping procedure used and which reported results as new, original findings.

### Inclusion and Exclusion Procedure

#### Title and Abstract Review

We excluded studies in which the titles and abstracts did not refer to tuberculosis or did not include the word “household” in reference to household contacts. If it was unclear whether a publication met these exclusion criteria, it was retained for full-text review.

#### Full-Text Review I

Full-text PDFs were downloaded and converted to text files. The ‘*grep*’ (‘global regular expression print’) command line tool was used to search for the following molecular typing keywords in each full-text document: “WGS”, “whole genome sequencing”, “spoligotyping”, “spoligotype”, “Spacer oligonucleotide typing”, “PCR”, “RFLP”, “restriction fragment length polymorphism”, “sequenced”, “typed”, “molecular typing”, “MIRU”, and “VNTR”. The “*-i*” option was applied accordingly in order to specify non-case-sensitive searches where appropriate. Publications which returned positive search results for one or more of the identified molecular typing terms were included; publications which returned negative search results for all keywords were excluded.

Each qualifying publication from the ‘*grep*’ search was manually read and closely inspected to evaluate whether the use of the molecular typing keywords aligned with our stated study definition. Publications which were found upon manual inspection to be commentaries, textbook excerpts, or otherwise not household-based designs were excluded. Publications where molecular typing terms only appeared in the reference list or that clearly did not genotype both index and household contact patients were excluded.

#### Full-Text Review II

The remaining full-text PDFs of publications found to have satisfied the inclusion and exclusion criteria of the Title and Abstract Review as well as Full-Text Review I were read to determine whether they fulfilled the study definition. Publications which systematically identified index TB patients and their household patients, included more than five households, genotyped household patients to compare them with their index patients, provided information about this household concordance in the paper or its supplement, and indicated the sequencing method used were included. Publications which failed to satisfy one or more aspects of this description were excluded.

---

**Table A1.** Lima study patient characteristics. Index and HHC characteristics of pairs are stratified by the presence of genetic linkage.

|  | N | Linked | Not Linked | p-value |
| --- | --- | --- | --- | --- |
| Index age |  |  |  | 0.823 |
| ≤ 35 | 77 | 53 (69%) | 24 (31%) |  |
| >35 | 32 | 21 (66%) | 11 (34%) |  |
| Index gender |  |  |  | 0.098 |
| Female | 46 | 27 (59%) | 19 (41%) |  |
| Male | 63 | 47 (75%) | 16 (25%) |  |
| Index HIV status |  |  |  | 0.241 |
| No | 106 | 73 (69%) | 33 (31%) |  |
| Yes | 3 | 1 (33%) | 2 (67%) |  |
| Index diabetes status |  |  |  | 1 |
| No | 105 | 70 (67%) | 35 (33%) |  |
| Yes | 1 | 1 (100%) | 0 (0%) |  |
| Index smear grade |  |  |  | 0.976 |
| Negative | 28 | 19 (68%) | 9 (32%) |  |
| + | 31 | 21 (68%) | 10 (32%) |  |
| ++ | 25 | 16 (64%) | 9 (36%) |  |
| +++ | 23 | 16 (70%) | 7 (30%) |  |
| Index cavitory disease |  |  |  | 0.663 |
| No | 72 | 50 (69%) | 22 (31%) |  |
| Yes | 36 | 23 (64%) | 13 (36%) |  |
| Index drug resistant profiles |  |  |  | 0.523 |
| DS | 59 | 42 (71%) | 17 (29%) |  |
| DR | 44 | 28 (64%) | 16 (36%) |  |
| HHC age |  |  |  | 0.465 |
| Children | 25 | 19 (76%) | 6 (24%) |  |
| ≥16 | 84 | 55 (65%) | 29 (35%) |  |
| HHC gender |  |  |  | 0.008 |
| Female | 61 | 48 (79%) | 13 (21%) |  |
| Male | 48 | 26 (54%) | 22 (46%) |  |
| HHC diabetes status |  |  |  | 0.549 |
| No | 105 | 70 (67%) | 35 (33%) |  |
| Yes | 3 | 3 (100%) | 0 (0%) |  |
| HHC BCG scars |  |  |  | 0.773 |
| 0 | 105 | 10 (62%) | 6 (38%) |  |
| ≥1 | 3 | 64 (69%) | 29 (31%) |  |

**Table A2.** Systematic Review Search Strategy.

| Database | Search Term | Filters |
| --- | --- | --- |
| PubMed | ((((("Tuberculosis"[MeSH] OR "Mycobacterium tuberculosis"[MeSH] OR "tuberculosis"[TI]))) AND (("contact investigation" OR "contact screening" OR "contact examination" OR "household screening" OR "contact screen" OR Contact Tracing[Mesh])) AND (("close family contact" OR "family contact" OR "close contact" OR "household" OR "tuberculosis contact" OR "childhood contact" OR "child contact" OR "adult contact" OR "contact")))) | *Publication date:*<br>2006/1/1 -<br>2025/1/21 |
| Web of Science | *ALL=((((("Tuberculosis" OR "Mycobacterium tuberculosis" OR "tuberculosis")) AND ("contact investigation" OR "contact screening" OR "contact examination" OR "household screening" OR "contact screen" OR Contact Tracing)) AND ("close family contact" OR "family contact" OR "close contact" OR "tuberculosis contact" OR "household" OR "childhood contact" OR "child contact" OR "adult contact" OR "contact"))))* | *Publication date:*<br>2006/1/1 -<br>2025/1/21 |
| Embase | ('tuberculosis'/exp OR 'mycobacterium tuberculosis' OR tuberculosis OR 'tuberculosis transmission'/exp OR 'tuberculosis transmission') AND ('contact investigation' OR 'contact screening' OR 'contact examination' OR 'household screening' OR 'contact screen') AND ('close family contact' OR 'family contact' OR 'close contact' OR 'household contact' OR 'household contact' OR 'household infection' OR 'tuberculosis contact' OR 'household transmission' OR 'childhood contact' OR 'adult contact' OR 'contact') AND [2006-2023]/py | *Publication date:*<br>2006-2025 |

**Table A3.** Quality Assessment.

| Study | 1. Were patients recruited by a systematic sample or census? | 2. Were at least 75% of identified eligible index patients enrolled? | 3. Was an appropriate index patients and household definition used in the study? | 4. Was there an initial screening of all HHCs for TB disease? | 5. Is the reported matching proportion restricted to patients with pulmonary TB? | 6. Was the time between index and contact patients diagnosis constrained to within 2 years? | 7. Was there active follow-up of HHCs to find secondary TB patients? | 8. Were at least 75% of culture-positive index-contact patients pairs genotyped? | Score |
| --- | --- | --- | --- | --- | --- | --- | --- | --- | --- |
| Marais et al. | Yes | Yes | Yes | Yes (study team) | Yes | Yes | No | Yes | 7 |
| Sia et al. | Yes | Not Reported | Yes | Yes (previous study team) | Yes | No | No | Yes | 5 |
| Crampin et al. | Yes | Yes | Yes | Not Reported | Yes | No | No | Yes | 5 |
| Buu et al. | Yes | Yes | Yes | Not Reported | Yes | Yes | No | Yes | 6 |
| Whalen et al. | Yes | Not Reported | Yes | Yes (study team) | Yes | Yes | Yes | Yes | 7 |
| Becerra et al. | Yes | Not Reported | Yes | Yes (national program, notes) | Yes | No | No | Yes | 5 |
| Leung et al. | Yes | Yes | Yes | Yes (local program) | Yes | No | No | Yes | 6 |
| Conceição et al. | Not Reported | Not Reported | Yes | Not Reported | Yes | No | Not Reported | Yes | 3 |
| Colangeli et al. | Yes | Yes | No | Yes (study team) | Yes | No | No | Yes | 5 |
| Morcillo et al. | No | Not Reported | Yes | Yes (national program) | No | No | No | Yes | 3 |
| Augustynowicz-Kope et al. | No | Not Reported | Yes | Not Reported | Yes | Yes | No | Yes | 4 |
| Borrell et al. | Yes | Yes | Yes | Yes (study team) | No | Yes | No | No | 5 |
| Lalor et al. | Yes | Yes | Yes | Yes (national program) | No | Yes | No | Yes | 6 |
| Our study in Lima | Yes | Not Reported | Yes | Yes | Yes | Yes | Yes | Yes | 7 |

**Table A4.** Genetic Matching by Household Contact Age.

| Study | Genotyping Method | Number of<br>Matched Index-Child Pairs | Number of Total<br>Index-Child<br>Pairs | Child HHC<br>Matching<br>Proportion | Number of<br>Matched Index-Adult Pairs | Number of Total<br>Index-Adult<br>Pairs | Adult HHC<br>Matching<br>Proportion |
| --- | --- | --- | --- | --- | --- | --- | --- |
| Sia et al | Spoligotyping | 0 | 1 | 0% | 11 | 15 | 73% |
|  | MIRU-VNTR (12) | 0 | 1 | 0% | 12 | 15 | 80% |
|  | IS6110 RFLP | 0 | 1 | 0% | 6 | 15 | 40% |
| Whalen | IS6110 RFLP | 26 | 29 | 90% | 20 | 32 | 63% |
| Conceicao et al <sup>a</sup> | MIRU-VNTR (24) | 2 | 3 | 67% | 26 | 31 | 83% |
| Augustyowicz-Kopec et al <sup>b</sup> | Spoligotyping | 1 | 1 | 100% | 37 | 42 | 88% |
|  | MIRU-VNTR (15) | 1 | 1 | 100% | 30 | 42 | 71% |

<sup>a</sup> One household (HHC22) was missing age information and was excluded
<sup>b</sup> 3 additional children were indicated to be index cases, with 2 of 3 genetically matching their household contacts

**Table A5.** Genetic Matching by Index and Household Contact Gender.

| Study | Genotyping Method | Female IP Matching | Male IP Matching | Female HHC Matching | Male HHC Matching |
| --- | --- | --- | --- | --- | --- |
| Sia et al | Spoligotyping | 3/3 = 100% | 8/13 = 62% | 4/5 = 80% | 7/11 = 64% |
|  | MIRU-VNTR (12) | 2/3 = 67% | 10/13 = 77% | 5/5 = 100% | 7/11 = 64% |
|  | IS6110 RFLP | 1/3 = 33% | 5/13 = 38% | 3/5 = 60% | 3/11 = 27% |
| Augustyowicz-Kopec et al <sup>a</sup> | Spoligotyping | 11/14 = 78% | 26/28 = 93% | 13/15 = 87% | 24/27 = 89% |
|  | MIRU-VNTR (15) | 10/14 = 71% | 20/28 = 71% | 11/15 = 73% | 19/27 = 70% |
| Buu et al | Spoligotyping | Not reported | Not reported | 1/2 = 50% | 4/10 = 40% |
|  | MIRU-VNTR (15) |  |  | 0/2 = 0% | 2/10 = 20% |
|  | IS6110 RFLP |  |  | 0/1 = 0% | 1/4 = 25% |
| Conceicao et al <sup>b</sup> | MIRU-VNTR (24) | 10/11 = 91% | 18/23 = 78% | 15/17 = 88% | 13/16 = 81% |

Definitions: IP, index patient; HHC, household contact
<sup>a</sup> One mixed-gender household (AE) did not indicate an index case and was excluded
<sup>b</sup> One household (HHC22) was missing gender information and was excluded

## References

1. Harries AD, Kumar AM, Satyanarayana S, Thekkur P, Lin Y, Dlodlo RA, et al. The Growing Importance of Tuberculosis Preventive Therapy and How Research and Innovation Can Enhance Its Implementation on the Ground. Tropical Medicine and Infectious Disease. 2020;5(2).

2. Salazar-Austin N, Mulder C, Hoddinott G, Ryckman T, Hanrahan C, Velen K, et al. Preventive Treatment for Household Contacts of Drug-Susceptible Tuberculosis Patients. Pathogens. 2022;11(11).

3. Martinez L, Shen Y, Mupere E, Kizza A, Hill PC, Whalen CC. Transmission of Mycobacterium Tuberculosis in Households and the Community: A Systematic Review and Meta-Analysis. American Journal of Epidemiology. 2017;185(12).

4. Sagili K, Muniyandi M, Shringarpure K, Singh K, Kirubakaran R, Rao R, et al. Strategies to detect and manage latent tuberculosis infection among household contacts of pulmonary TB patients in high TB burden countries - a systematic review and meta-analysis. Tropical medicine & international health. 2022;27(10).

5. Becerra MC, Huang C-C, Lecca L, Bayona J, Contreras C, Calderon R, et al. Transmissibility and potential for disease progression of drug resistant Mycobacterium tuberculosis: prospective cohort study. BMJ. 2019;367.

6. McIntosh A, Jenkins H, Horsburgh C, Jones-López E, Whalen C, Gaeddert M, et al. Partitioning the risk of tuberculosis transmission in household contact studies. PloS one. 2019;14(10).

7. McIntosh A, Doros G, Jones-López E, Gaeddert M, Jenkins H, Marques-Rodrigues P, et al. Extensions to Bayesian generalized linear mixed effects models for household tuberculosis transmission. Stat Med. 2017;36(16).

8. Li H, Durbin R. Fast and accurate long-read alignment with Burrows–Wheeler transform. Bioinformatics. 2010;26(5).

9. Li H, Handsaker B, Wysoker A, Fennell T, Ruan J, Homer N, et al. The Sequence Alignment/Map format and SAMtools. Bioinformatics. 2009;25(16).

10. Walker BJ, Abeel T, Shea T, Priest M, Abouelliel A, Sakthikumar S, et al. Pilon: An Integrated Tool for Comprehensive Microbial Variant Detection and Genome Assembly Improvement. PLOS ONE. 2014;9(11).

11. Comas I, Chakravartti J, Small PM, Galagan J, Niemann S, Kremer K, et al. Human T cell epitopes of Mycobacterium tuberculosis are evolutionarily hyperconserved. Nature Genetics 2010 42:6. 2010;42(6).

12. Marin M, Vargas R, Harris M, Jeffrey B, Epperson L, Durbin D, et al. Benchmarking the empirical accuracy of short-read sequencing across the M. tuberculosis genome. Bioinformatics. 2022;38(7).

13. Coll F, McNerney R, Guerra-Assunção JA, Glynn JR, Perdigão J, Viveiros M, et al. A robust SNP barcode for typing Mycobacterium tuberculosis complex strains. Nature Communications 2014 5:1. 2014;5(1).

14. Trauner A, Liu Q, Via L, Liu X, Ruan X, Liang L, et al. The within-host population dynamics of Mycobacterium tuberculosis vary with treatment efficacy. Genome biology. 2017;18(1).

15. WHO Stop TB Partnership CTS. Guidance for National Tuberculosis Programmes on the management of tuberculosis in children. Int J Tuberc Lung Dis. 2006.

16. National Guidelines on Management of Tuberculosis in Children. Kenyan Ministry of Health, Division of Leprosy, Tuberculosis and Lung Disease; 2013.

17. Martinez L, Lo NC, Cords O, Hill PC, Khan P, Hatherill M, et al. Paediatric tuberculosis transmission outside the household: challenging historical paradigms to inform future public health strategies. The Lancet Respiratory Medicine. 2019;7(6).

18. Supply P, Allix C, Lesjean S, Cardoso-Oelemann M, Rüsch-Gerdes S, Willery E, et al. Proposal for Standardization of Optimized Mycobacterial Interspersed Repetitive Unit-Variable-Number Tandem Repeat Typing of Mycobacterium tuberculosis. J Clin Microbiol. 2006;44(12).

19. WHO TB burden estimates. Global Tuberculosis Programme: World Health Organization; 2024.

20. Colangeli R, Gupta A, Vinhas SA, Chippada Venkata UD, Kim S, Grady C, et al. Mycobacterium tuberculosis progresses through two phases of latent infection in humans. Nature Communications. 2020;11(1).

21. Marais BJ, Hesseling AC, Schaaf HS, Gie RP, Helden PDv, Warren RM. Mycobacterium tuberculosis Transmission Is Not Related to Household Genotype in a Setting of High Endemicity. J Clin Microbiol. 2009;47(5).

22. Sia IG, Buckwalter SP, Doerr KA, Lugos S, Kramer R, Orillaza-Chi R, et al. Genotypic characteristics of Mycobacterium tuberculosis isolated from household contacts of tuberculosis patients in the Philippines. BMC Infectious Diseases. 2013;13(1).

23. Crampin AC, Glynn JR, Traore H, Yates MD, Mwaungulu L, Mwenebabu M, et al. Tuberculosis Transmission Attributable to Close Contacts and HIV Status, Malawi. Emerg Infect Dis. 2006;12(5).

24. Buu T, van Soolingen D, Huyen M, Lan N, Quy H, Tiemersma E, et al. Tuberculosis acquired outside of households, rural Vietnam - PubMed. Emerg Infect Dis. 2010;16(9).

25. Whalen CC, Zalwango S, Chiunda A, Malone L, Eisenach K, Joloba M, et al. Secondary Attack Rate of Tuberculosis in Urban Households in Kampala, Uganda. PLOS ONE. 2011;6(2).

26. Becerra MC, Franke MF, Appleton SC, Joseph JK, Bayona J, Atwood SS, et al. Tuberculosis in Children Exposed at Home to Multidrug-resistant Tuberculosis. The Pediatric Infectious Disease Journal. 2013;32(2).

27. Leung ECC, Leung CC, Kam KM, Yew WW, Chang KC, Leung WM, et al. Transmission of multidrug-resistant and extensively drug-resistant tuberculosis in a metropolitan city. Eur Respir J. 2013;41(4).

28. Conceição EC, Guimarães AEdS, Lopes ML, Furlaneto IP, Rodrigues YC, Conceição MLd, et al. Analysis of potential household transmission events of tuberculosis in the city of Belem, Brazil. Tuberculosis. 2018;113.

29. Morcillo NS, Imperiale BR, Giulio ÁD, Zumárraga MJ, Takiff H, Cataldi ÁA. Fitness of drug resistant Mycobacterium tuberculosis and the impact on the transmission among household contacts. Tuberculosis. 2014;94(6).

30. Augustynowicz-Kopeć E, Jagielski T, Kozińska M, Kremer K, Soolingen Dv, Bielecki J, et al. Transmission of tuberculosis within family-households. Journal of Infection. 2012;64(6).

31. Borrell Sn, Español M, Orcau An, Tudó G, March F, Caylà JA, et al. Factors Associated with Differences between Conventional Contact Tracing and Molecular Epidemiology in Study of Tuberculosis Transmission and Analysis in the City of Barcelona, Spain. J Clin Microbiol. 2009;47(1).

32. Lalor MK, Anderson LF, Hamblion EL, Burkitt A, Davidson JA, Maguire H, et al. Recent household transmission of tuberculosis in England, 2010–2012: retrospective national cohort study combining epidemiological and molecular strain typing data. BMC Medicine. 2017;15(1).

33. Kato-Maeda M, Metcalfe JZ, Flores L. Genotyping of Mycobacterium tuberculosis: application in epidemiologic studies. Future microbiology. 2011;6(2).

34. Duong T, Brigden J, Simon Schaaf H, Garden F, Marais B, Anh Nguyen T, et al. A Meta-Analysis of Levofloxacin for Contacts of Multidrug-Resistant Tuberculosis. NEJM Evidence. 2025;4(1).

35. Reid M, Agbassi YJP, Arinaminpathy N, Bercasio A, Bhargava A, Bhargava M, et al. Scientific advances and the end of tuberculosis: a report from the Lancet Commission on Tuberculosis. The Lancet. 2023;402(10411).

36. Reich-Stiebert N, Froehlich L, Voltmer J-B, Reich-Stiebert N, Froehlich L, Voltmer J-B. Gendered Mental Labor: A Systematic Literature Review on the Cognitive Dimension of Unpaid Work Within the Household and Childcare. Sex Roles 2023 88:11. 2023-04-29;88(11).

37. OECD. Caregiving in Crisis: Gender inequality in paid and unpaid work during COVID-19. 2021.

38. Ervin J, Taouk Y, Alfonzo LF, Hewitt B, King T. Gender differences in the association between unpaid labour and mental health in employed adults: a systematic review. The Lancet Public Health. 2022;7(9).

39. Hyde E, Greene ME, Darmstadt GL. Time poverty: Obstacle to women’s human rights, health and sustainable development. Journal of Global Health. 2020;10(2).

